# Knowledge Attitude and Practice towards Cutaneous Leishmaniasis in Sodo District Southern Ethiopia

**DOI:** 10.1101/2022.08.29.22279270

**Authors:** Lina Gazu, Zerish Zethu Nkosi, Nigatu Kebede

## Abstract

**Background:** In Ethiopia, Cutaneous Leishmaniasis (CL) is a common infectious disease. However, existing knowledge on community awareness is scarce.

**Objective:** Objectives is to access knowledge, attitude, and practices about CL in Sodo District.

**Methods:** Primary quantitative method using a cross-sectional descriptive approach was applied. Data were collected from face-to-face interviews held with 423 households between January to April 2018. Knowledge, attitude, and practice scores were obtained by aggregating responses to questions. Levels of these outcomes were determined by dichotomizing the generated scores using their mean vales. Percentages, frequency and mean values were used to descriptively understand the data. Logistic regression was used to model the binary outcomes. R Package Version 3.02 was used to conduct the statistical analysis.

**Results:** Of 423 participants 263 (61.9%), 226 (53.4%), and 213 (50.4%) have satisfactory knowledge, favourable attitude, and good practice about leishmaniasis. Majority are unable to identify leishmaniasis and unaware of its cause and transmission. Sandfly is considered “important biting and blood-sucking insect” by 210 (49.6%) but knowledge on biting time and breeding place was unsatisfactory. Most believed CL can be treated, is a serious problem and have a disfiguring outcome. However, most (59.3%, n=251) prefer use of traditional medication. Concerning practice, 288 (68.1%) have bed nets but personal protective measures are rarely used.

**Conclusion:** The level of overall knowledge and attitudes and practice in the current study was not adequate. This finding implies that there is a need for intensified education regarding CL.

## Introduction

Leishmaniasis is a parasitic disease caused by flagellated protozoans of the Leishmania genus. It is spread by the bite of a female hematophagous sandfly vector, which is widespread in humans and certain animals (anthropozoonoses). Leishmaniasis is prevalent in nearly a hundred countries, resulting in millions of new cases and up to 65,000 deaths per year ([1]). Cutaneous Leishmaniasis (CL), mucosal involvement (MCL), and systemic visceral involvement (VL) are all clinical manifestations of the illness in humans (2).

CL is the most common type of leishmaniasis worldwide. CL affects over one million people worldwide per year, with hotspots in Afghanistan, Algeria, Iran, Pakistan, Peru, Brazil, Saudi Arabia, Colombia, and Tunisia (3). Due to environmental changes, host immune status treatment failure and drug resistance, re-emergency of CL is recently observed in many endemic countries (4).

In Ethiopia, the yearly burden of CL is estimated to be between 20,000 to 30,000 cases putting the country among high CL burden countries (5). *Leishmania aethiopica* is the most important cause of CL in Ethiopia (6). The interesting features of leishmanaisis caused by *L. aethiopica* are atypical lesions appearing commonly on the face, healing of lesion taking longer time and lesions are more severe in their presentation than that of other species (6).

The first CL case in Ethiopia was reported in the early 20th century. “Kunchir” in Gojam, “Finchoftu” in northern Shoa, Gonder, and sections of Wollo, “Chewie” in Sodo, “Shahegne” in north Shewa, “Volbo” in Ocholo, “Giziwa” in Tigray, and “Simbirahalkani” in Wollega are some of the local names for CL (7). CL is endemic in Ethiopian highlands with an altitude > 2,000 meters (8). CL has been estimated to threaten about 30 million Ethiopians, predominantly from the highlands of Amhara, Tigray, Oromia, and the Southern Nations, Nationalities, and Peoples’ Region (9).

Disfigurement from CL cause both social and psychological influences such as nervousness, tension and depression leading to low quality of life which in turn affectseconomic productivity of individuals (10). However, adding to the growing proof of an evolving challenge, no vaccines exist for human, and difficulty in treatment, particularly in resource-limited countries like Ethiopia (9). Data on precise level of CL is deficient in Ethiopia and the country has not designed a program to control the vectors transmitting the disease (11). Moreover, the disease is the most overlooked disease in Ethiopia among all tropical diseases neglected so far. The Ethiopian Federal Ministry of Health has established a foundation to construct a leishmaniasis control program, recognizing the growing burden of the disease. In order to control and prevent leishmaniasis in Ethiopia, the Ministry has also designated vector control, health education, social mobilisation, and information system strengthening as priorities (12). Before planning any awareness creation programme, it is vital to gather evidence on the level of existing knowledge and gaps in the community. This evidence will serve as an indicator for health education and promotion activities, identify research gaps, and feed data for planning applicable policies to monitor government and community attempts. Nevertheless, there are few KAP surveys in the country and none in the study area to the knowledge of the researcher. Therefore, a KAP survey in each endemic area will be vital to design intervention strategies against the disease. Putting this fact in to account the present study was designed to assess KAP of Sodo community towards CL.

## Methods

### Description of the study area

The research was carried out in the Sodo District. Sodo District is situated in Garage Zone of SNNP’s Regional State (Fig 1). The geographical location of the district according to Ethiopian Central Statistical Agency (CSA), is from 8° 09’ to 8° 45’ North latitude and from 38° 37’ to 38° 71’ East longitude (13). It is located approximately 103 kilometres south of Addis Ababa on the route. The Woreda has an area of 88,553.3 hectares. The Woreda also shared its boundary with Meskan Woreda on the south, on the west, north, and the east by Oromia Region.

**Fig 1.**
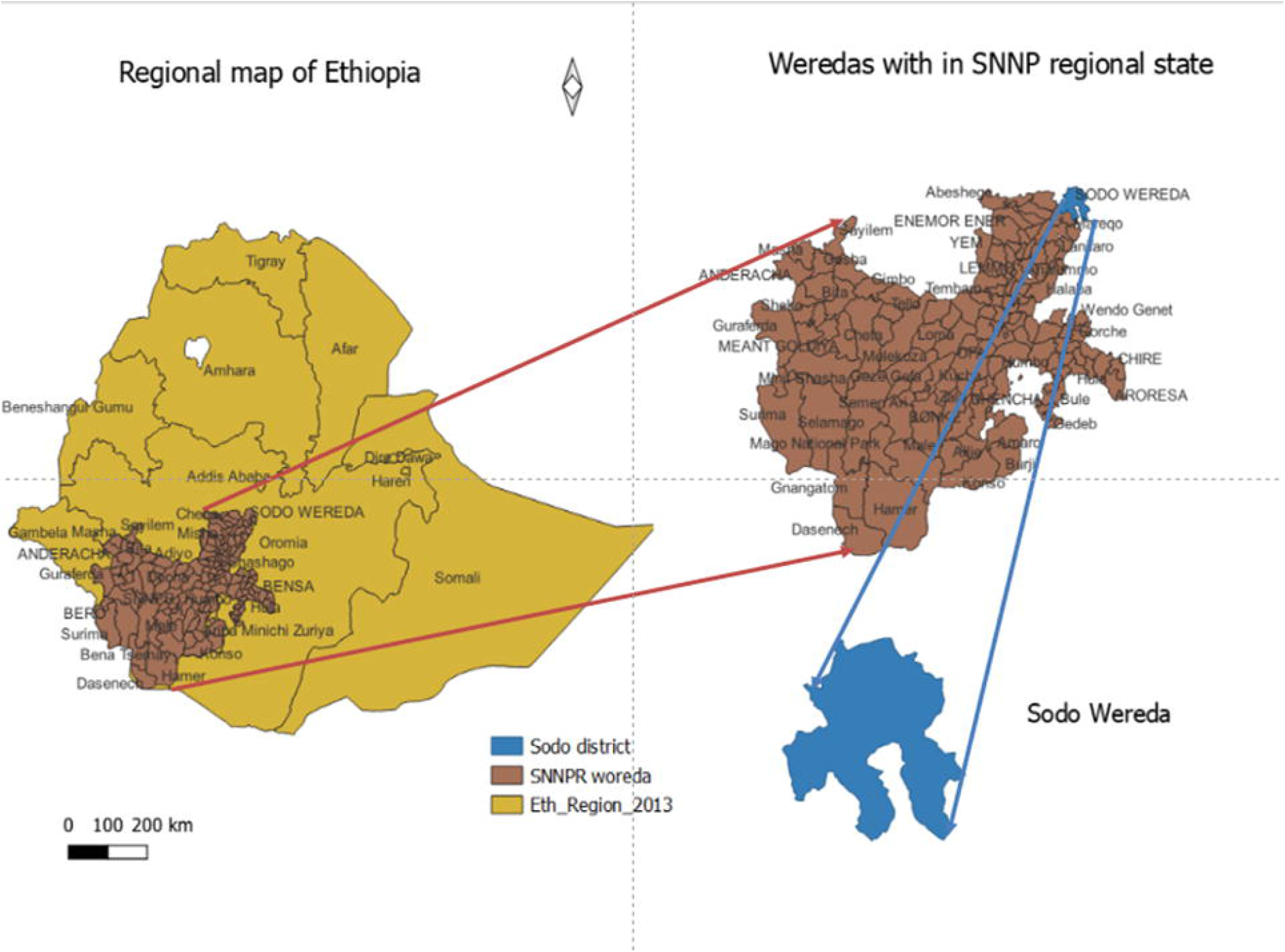
Map of study area. SNNPR (Southern Nation, Nationalities, and people Region – currently there are two additional regions namely Sidama and Southe West Ethiopia regions, which were created after the survey was conducted). The base layer dataset included shapefiles obtained from open AFRICA (https://africaopendata.org/dataset/ethiopia-shapfiles) which is licensed under a Creative Commons Attribution. The map was created using QGIS version 3.20.1.

The district is expected to have a total population of 175,725 people, with 89,619 (51%) women and 90% of the population residing in rural regions(14). The district is scheduled to have eight health centers, 55 health posts, two private clinics, and three pharmacies (15). Based on traditional healers, district health professionals, and secondary data six kebeles, namely, Kela Zuria, Kola Nurena, Michael Semero, Adazer, Beka, and Genete Mariam were selected with a total household of 2632. The number of households in each kebele is depicted in Table 1.

**Table 1:** Total number of households and population in selected kebeles.

| Keble | No of households | Population |
| --- | --- | --- |
| Kela Zuria | 333HH | 2028 |
| Kola Nurena | 362HH | 2389 |
| Michael Semero | 475HH | 3055 |
| Adazer | 610HH | 3477 |
| Beka | 299HH | 1678 |
| Genete Mariam | 553HH | 2912 |

### Sample and sampling methods

For determining the sample size for the KAP questionnaire study, 50% of the communities are assumed to be knowledgeable about CL with 2% marginal error and 95% confidence interval. The following single population proportion formula was employed to get the definite sample size with a 10% non-response rate.

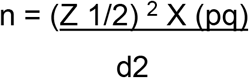

Where: d= Degree of marginal error (5%)

Z = level of confidence (95%)

P = CL prevalence (50%), q = 1-p

adding 10% non-response rate,

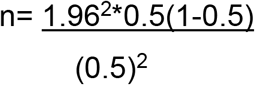

The required sample size was 423 households.

Dividing the total sample to each kebele was done by applying the following population proportion formula to sample 423 HH

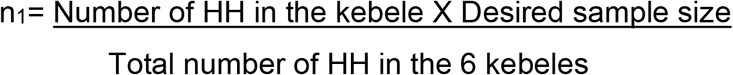

Therefore, 54,58, 76,98,48 and 89 questioners were administered in Kela Zoria, Kola Nurena, Semero, Adazer, Beka and Genete Mariam, respectively.

### Data collection

A standardized questionnaire (S1Text) was prepared and administered to each participant who agreed to be part of the study. The survey tool was translated to Amharic language keeping its original meaning. The household heads in the identified houses or another older family member, if the household head is not around, were interviewed. Data was collected by nine health extension workers, with a diploma and bachelor’s degrees, were selected from each kebele of the Sodo District for this purpose. The data collectors received a one-day training which took one full day before the beginning of data gathering.

### Data quality assurance

A standardized questionnaire was prepared, and it was translated into local language (Amharic) and the data collectors used this version to interview study participants. The questionnaire was pre-tested to maximize the quality of data by finding out how well the questions are understood by the interviewees. For this purpose, 30 individuals were interviewed at the stage of pre-testing. During the actual data collection, the completed questionnaire was checked for completeness daily by the investigator.

### Ethical consideration

The research was approved by the Ethics Committee of the University of South Africa’s (UNISA) Department of Health Studies (Reference number:REC-012714-039/HSHDC/457/2015). The SNNPR health bureau, Gurage zone administration, and Sodo Health office all issued approval letters. Written consent was obtained from study participants.

### Inclusion and exclusion criteria

Participants comprised any consenting individuals residing in the selected six kebeles. Individuals who live outside the defined areas were excluded. Households that have been visited three times without finding an adult to approach for consent were considered non-responsive, excluded from the study, and replaced with an alternative household.

### Data analysis

Knowledge was measured using an 8-item questionnaire containing: If they could name, the disease after being shown a picture of CL manifestation, have you ever heard of CL? What is the cause of CL? What are major symptoms of CL? How is the disease transmitted? Whether they know the importance of the vector as a transmitter of leishmaniasis after being shown a picture of phletomine sandfly believed to exist in area, where do vectors breed? When does the vector prefer to bite?

The attitude was evaluated by using a 6-point questionnaire comprising: Do you think chewie can be treated? What is your preferred choice of medication for the treatment of Chewie? What is your opinion on the outcome of CL if left untreated? Do you feel that you are well informed about chewie? Willing to participate in CL control activities? Do you consider CL a serious health problem in your localities?

Prevention practice had 8 items including: How was Chewie treated? Use of Preventable action for CL? Type of methods used to prevent CL, having bed nets in the house, sleeping condition, using bed nets when sleeping, using repellents, and working time preference at peak temperature.

In obtaining measures for the outcomes a score of one was given for correct responses for each question and a score of zero was used for incorrect responses and ‘do not know’ responses. Knowledge, attitude and practice scores were then generated by adding responses given to these questions. Finally, total score was summarized taking the mean score of the composite variable as a cut-off so that anyone whose score is above the mean was classified as having satisfactory knowledge, favorable attitude or good practice on the above specific outcomes and those scoring below the cut-off value (below the mean score) were considered as having unsatisfactory knowledge, unfavorable attitude or poor practice for each outcome in each component.

Percentage, frequency, and mean values were used to descriptively summarise the data. To investigate independent variables that could potentially predict the probability of a higher level of overall knowledge, attitude, and practice of study participants about CL, a binary logistic regression analysis (both bivariate with crude odds ratio and multivariate with adjusted odds ratio) at 95% confidence intervals (CI) was performed. After calculating crude odds ratio and adjusted odds ratio, significant level was set if p-value is less than 0.05. Tables as well as bar graphs were used to present results and data were computerised and analysed using R Package Version 3.02.

## Results

### Socio-demographic characteristics

A total of 423 individuals were involved in this study; 219 (51.8%) of the participants were males and 204 (48.2%) were females. One hundred fifty (35.5%) were in the age range of 34.5 to 44.5 years and 107 (25.3%) are in the age group of 24.5–34.5. Orthodox 391 (92.4%) was the dominant religion in Sodo District. Participant’s major occupation was farming [216 (51.1%)]. The average family size within the study participants was 5.76 and the average duration of stay in the district was 32.79 years. Accordingly, only a few proportions of the study participants 81 (19.1%) came from another place while the rest 342 (80.9%) are originally from the same district. The general socio-demographic features of the respondents were presented in Table 2.

**Table 2:** Demographic characteristics of the respondents (N=423).

| Characteristics | Frequency | Percent |
| --- | --- | --- |
| **Gender** |  |  |
| Male | 219 | 51.8 |
| Female | 204 | 48.2 |
| **Age** |  |  |
| 18.0-24.5 | 24 | 5.7 |
| 24.5–34.5 | 107 | 25.3 |
| 34.5- 44.5 | 150 | 35.5 |
| 44.5-54.5 | 82 | 19.4 |
| Over 54.5 | 60 | 14.2 |
| **Religion** |  |  |
| Orthodox | 391 | 92.4 |
| Protestant | 29 | 6.9 |
| Muslim | 3 | 0.7 |
| **Educational level** |  |  |
| Illiterate (cannot read & write) | 216 | 51.1 |
| Can read and write (Only) | 119 | 28.1 |
| Primary education (1-6 grade) | 63 | 14.9 |
| Secondary education (8-12 grade) & above | 25 | 5.9 |
| **Occupation** |  |  |
| Farmer | 216 | 51.1 |
| Housewives | 188 | 44.4 |
| Trader | 19 | 4.5 |
| **If yes location of farmland?** |  |  |
| Near home | 218 | 51.5 |
| Near a river | 27 | 6.4 |
| Both | 106 | 25.1 |
| other | 72 | 17.0 |
| **Place of birth** |  |  |
| Same district | 342 | 80.9 |
| Another place | 81 | 19.1 |

Considering the education level of the respondents, 216 (51.1%) were illiterates while 119 (28.1%) of the participants can read and write only. None of them travelled out of their district in six months of the study time. Of the 351 (82.8%) participants who claim to have farming land, the majority 218 (51.5%) confirmed the land is situated near their home.

### Knowledge of the community about cutaneous leishmaniasis

Knowledge of CL among study participants is presented in Table 3. Two hundred and sixteen (51.1%) of the participants were able to identify the disease as CL after they were shown a picture illustrating cases of CL reported from different parts of Ethiopia (S2 Text) and 350 (82.7%) of the participants have heard about CL. However, the majority of 350 (82.7%) of the participants have heard about CL mostly being informed by families, friends, neighbours, and colleagues 286 (67.5%) and from being exposed to the disease 38 (9.0%).

**Table 3:**
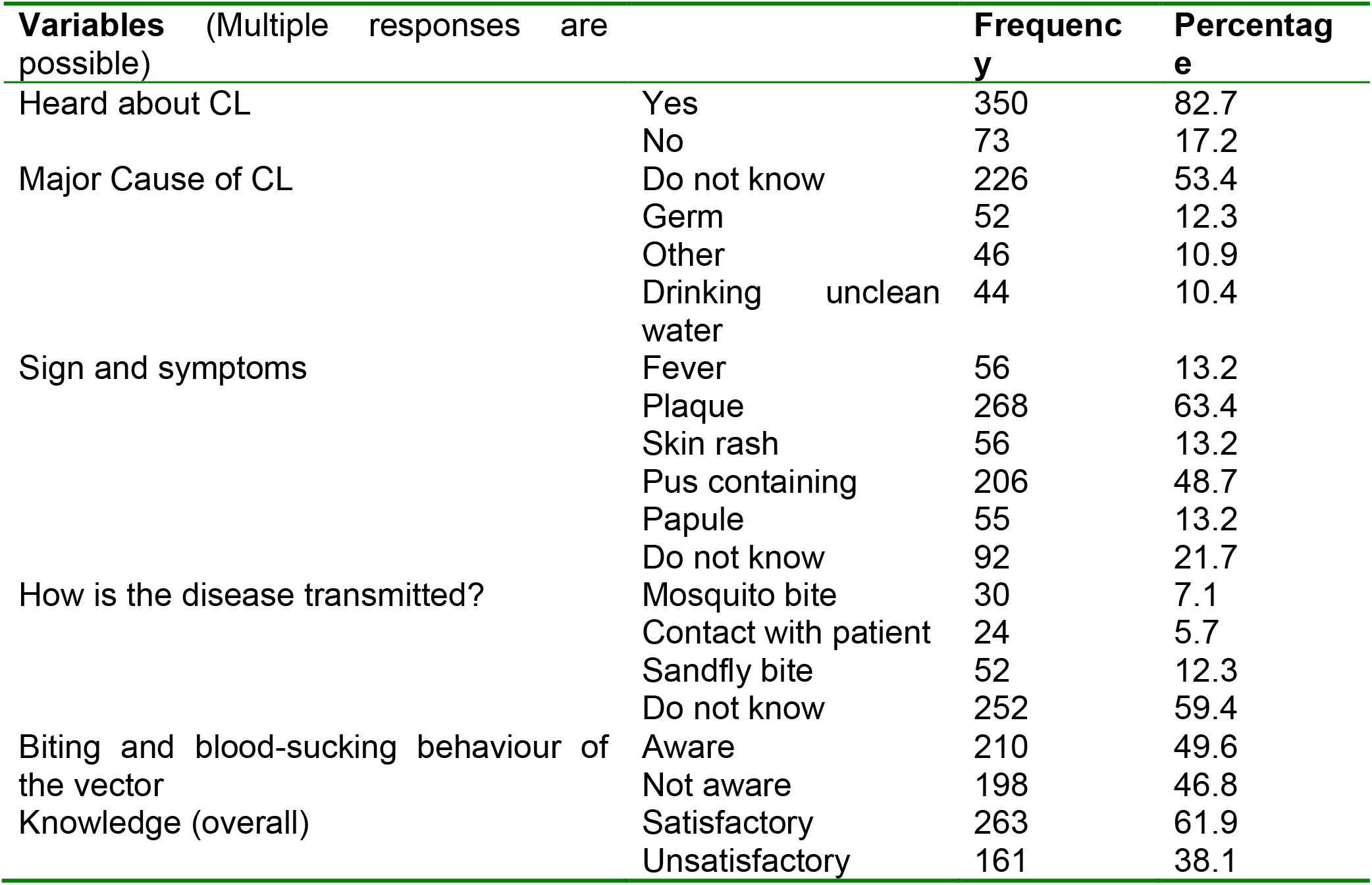
Knowledge of CL among study participants (N=423)

When asked if they know anybody who is infected by the disease in their area, most 301 (71.2%) of the respondents knew someone infected with CL. Of these, 218 (51.5%) knew that the person they know to have CL was infected in the past and 80 (18.9%) claim that the person they know is actively infected. The individuals they know of being infected are neighbours, family members, cousin, and self in 154 (36.4%), 52 (12.3%), 25 (5.9%), and 38 (9.0%). of the respondents respectively (Fig 2).

**Fig 2:**
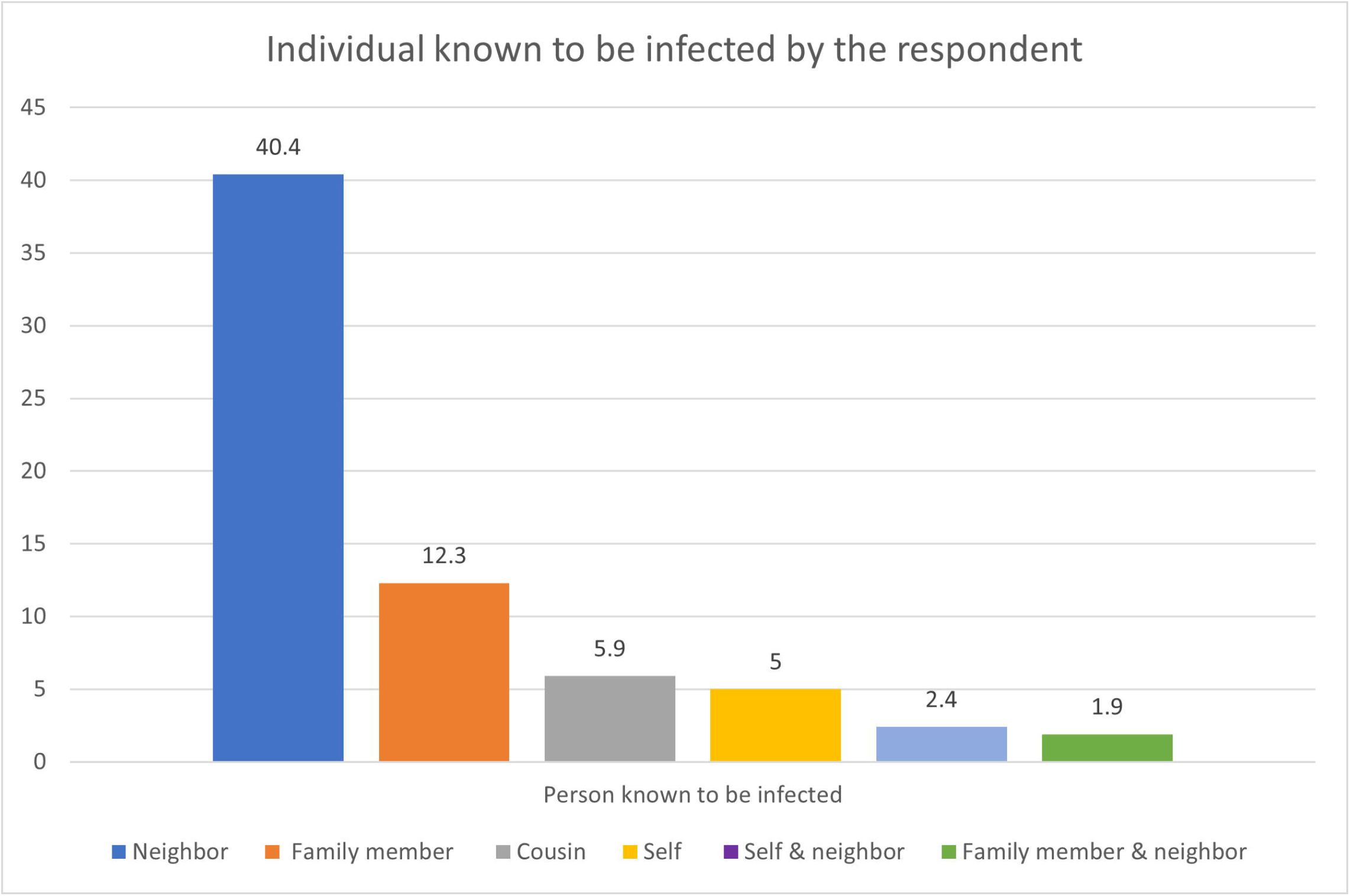
Relationship of persons known to be infected with CL and respondent. Two hundred six (53.4%) of the respondents claim that they do not know the cause of CL, 52 (12.3%) mentioned germ, and 44 (10.4%) say drinking unclean water could cause the disease. More than half 252 (59.4%) of the study participants are not aware on how the disease is transmitted while only a small proportion of the study population (52 (12.35%)) knew the disease is communicated by a bite of sandfly vector. The current study revealed that plaque and pus-containing wounds were mentioned as the main clinical symptoms of CL by 268 (63.4%) and 206 (48.6%) of the respondents respectively while 92 (21.7%) admitted that they did not know any symptom of CL.

Nose (316 (74.5%)), face (305 (71.9%)), forehead (116 (27.4%)) and ear (102 (24.1%)) are parts of the body mentioned by the respondents to be mostly infected by the lesion (called “kuselet” in Amharic) from CL infection. In this study, only 93 (21.9%) knew that there is a possibility of acquiring leishmaniasis by travelling to areas that are prone to the disease. It is obvious that these parts of the body are usually left uncovered thus are easily assessable for vectors

To measure the knowledge of study participants about the sandfly vector, a picture of the vector that is believed to exist in the southern part of Ethiopia was shown to the respondents and their reply was recorded (S3 Text). As a result, nearly half 210 (49.6%) reported to know the biting and blood-sucking behaviour of phletomine sandfly. In the current study, 123 (29.0%) and 139 (32.8%) of the participants do not know the breeding place and the preferred biting time of the vector, respectively. But 178 (42%) said that the vector bits mostly at dusk. Dirty places 218 (51.5%), water ponds 194 (45.9%), and garbage collection sites 111 (26.2%) are the most mentioned breeding sites of the vector. A little percentage of response was given to cervices in the house 18 (4.3%), thatched roof 17 (4.0%), and cattle shades 20 (4.7%).

### Factors affecting knowledge of the study participants

The result of the bi-variate analysis reveals a statistically significant relationship between gender (p< 0.000), education level (p< 0.002 & p< 0.011), occupation (p< 0.000), place of birth (P< 0.026), and knowledge of a person with CL (p< 0.000) and CL knowledge of the participant. Hence, females are knowledgeable than males [COR=2.9; 95% CI, 1.919-4.371]. Accordingly, females are three times likely to have satisfactory knowledge than males (Table 4).

**Table 4:** Comparison of demographic variables with knowledge of CL (N=423).

| Variables |  | COR (95% CI) | AOR (95%CI) |
| --- | --- | --- | --- |
| **Gender** |  |  |  |
|  | Male | 1.00 | 1.00 |
|  | Female | 2.897(1.919-4.371) | 1.997(0.640-6.233) |
| **Age** |  |  |  |
|  | 18.0-24.5 | 1.00 | 1.00 |
|  | 24.5–34.5 | 1.345(0.535-3.385) | 0.951(0.339-2.666) |
|  | 34.5- 44.5 | 1.007(0.414-2.453) | 0.875(0.317-2.416) |
|  | 44.5-54.5 | 0.987(0.386-2.525) | 1.077(0.367-3.157) |
|  | Over 54.5 | 0.525(0.199-1.384) | 0.656(0.212-2.030) |
| **Educational level** |  |  |  |
|  | Illiterate | 1.00 | 1.00 |
|  | Able to write & read | 0.489(0.310-0.771) | 0.838(0.470-1.494) |
|  | Primary | 2.515(1.238-5.105) | 3.879(1.729-8.703) |
|  | Secondary & above | 0.491(0.213-1.130) | 0.613(0.238-1.580) |
| **Occupation** |  |  |  |
|  | Farmer | 1.00 | 1.00 |
|  | Trader | 1.683(0.638-4.437) | 1.039(0.296-3.645) |
|  | Housewife | 2.945(1.926-4.502) | 1.527(0.477-4.891) |
| **Place of birth** |  |  |  |
|  | Same district | 1.00 | 1.00 |
|  | Another place | 1.836(1.075-3.136) | 0.750(0.369-1.524) |
| **Know a person with CL infection** |  |  |  |
|  | Yes | 1.00 | 1.00 |
|  | No | 0.156(0.098-0.248) | 0.163(0.099-0.268) |

Respondents who can read and write are less likely to be knowledgeable compared to those who are illiterate [COR=0.49; 95% CI, 0.310-0.771]. This means there is no significant difference between illiterates and who can read and write for acquiring knowledge of CL. On the contrary, respondents who receive primary education are more likely to have a better knowledge of CL than those who are illiterate [COR=2.5; 95% CI, 1.238-5.105].

Individuals who are not originally from the Sodo District are knowledgeable compared to participants who come from other places [COR=1.84; 95% CI, 1.075-3.136]. Study participants who claim not to know a person infected with the disease are less knowledgeable than those who do [COR=0.156; 95% CI, 0.098-0.248].

Multivariate analysis also shows a statistically significant association between educational level, previous knowledge of a person with CL and level of knowledge. Respondents receiving primary education [AOR=3.879; 95% CI, 1.729-8.703] are found to be knowledgeable than respondents who are illiterates. This shows that for education to be a factor affecting knowledge, the level must be at least to receiving a primary level education. Those who do not know someone infected with CL are less knowledgeable than those who do [AOR=0.163; 95% CI, 0.099-0.268]. From the results, it can generally be concluded that after adjusting for all variables, receiving primary education, and knowing someone infected with CL are factors that truly contribute to satisfactory knowledge of CL.

### Attitude towards cutaneous leishmaniasis in Sodo District

The attitude of the study participants in the area is recorded in Table 5. Two hundred and sixty-one (61.7%) of the participants in this study believed that CL can be treated. However, 70 (16.5%) of the members perceived that CL is not curable and 85 (20.1%) have no idea whether the disease can be treated or not. Partly 257 (60.6%) of the participants in this study share the fact that environmental hygiene is important for the prevention of CL transmission.

**Table 5:**
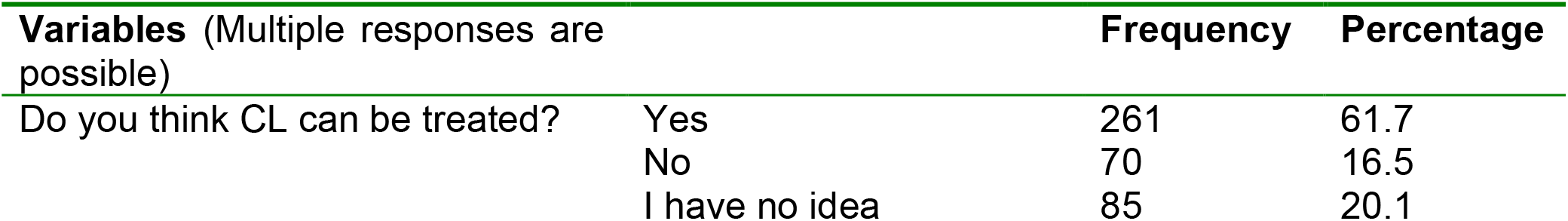
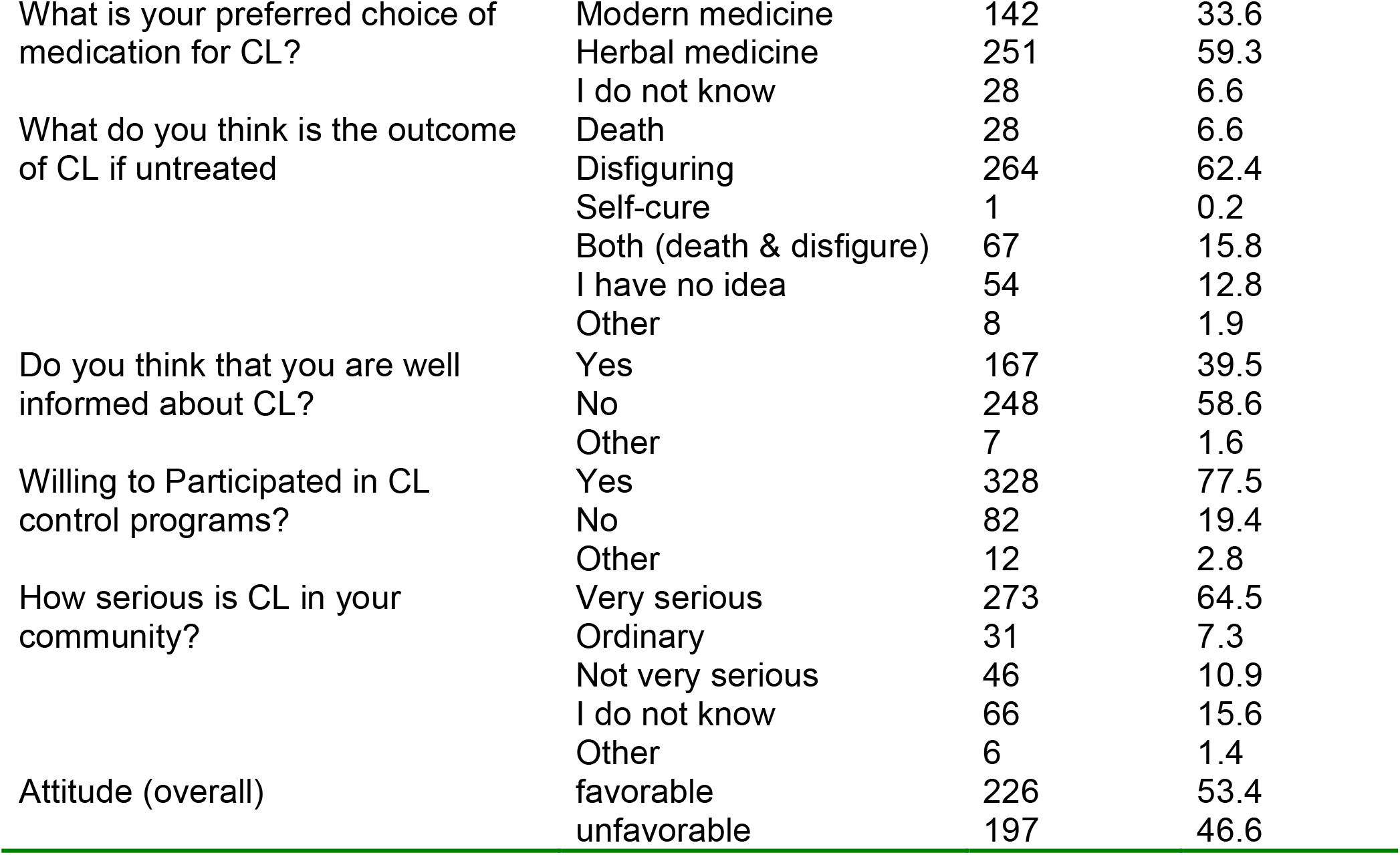
Attitude of the Sodo community towards CL (N=423).

| Variables (Multiple responses are possible) |  | Frequency | Percentage |
| --- | --- | --- | --- |
| Do you think CL can be treated? | Yes | 261 | 61.7 |
|  | No | 70 | 16.5 |
|  | I have no idea | 85 | 20.1 |
| What is your preferred choice of medication for CL? | Modern medicine | 142 | 33.6 |
|  | Herbal medicine | 251 | 59.3 |
|  | I do not know | 28 | 6.6 |
| What do you think is the outcome of CL if untreated | Death | 28 | 6.6 |
|  | Disfiguring | 264 | 62.4 |
|  | Self-cure | 1 | 0.2 |
|  | Both (death & disfigure) | 67 | 15.8 |
|  | I have no idea | 54 | 12.8 |
|  | Other | 8 | 1.9 |
| Do you think that you are well informed about CL? | Yes | 167 | 39.5 |
|  | No | 248 | 58.6 |
|  | Other | 7 | 1.6 |
| Willing to Participated in CL control programs? | Yes | 328 | 77.5 |
|  | No | 82 | 19.4 |
|  | Other | 12 | 2.8 |
| How serious is CL in your community? | Very serious | 273 | 64.5 |
|  | Ordinary | 31 | 7.3 |
|  | Not very serious | 46 | 10.9 |
|  | I do not know | 66 | 15.6 |
|  | Other | 6 | 1.4 |
| Attitude (overall) | favorable | 226 | 53.4 |
|  | unfavorable | 197 | 46.6 |

More than half 251 (59.3%) of the respondents preferred indigenous medication through the application of herbs by traditional healers. In the current study, of those who do not seek modern medication claim lack of trust in modern medication 165 (38.9%), trusting herbal medication 143 (33.7%) high cost of modern medication 95 (22.4%) and not having time to attend medical care because of being busy with work 69 (16.3%) as the main reasons for not seeking modern treatment (Fig 3).

**Fig 3:**
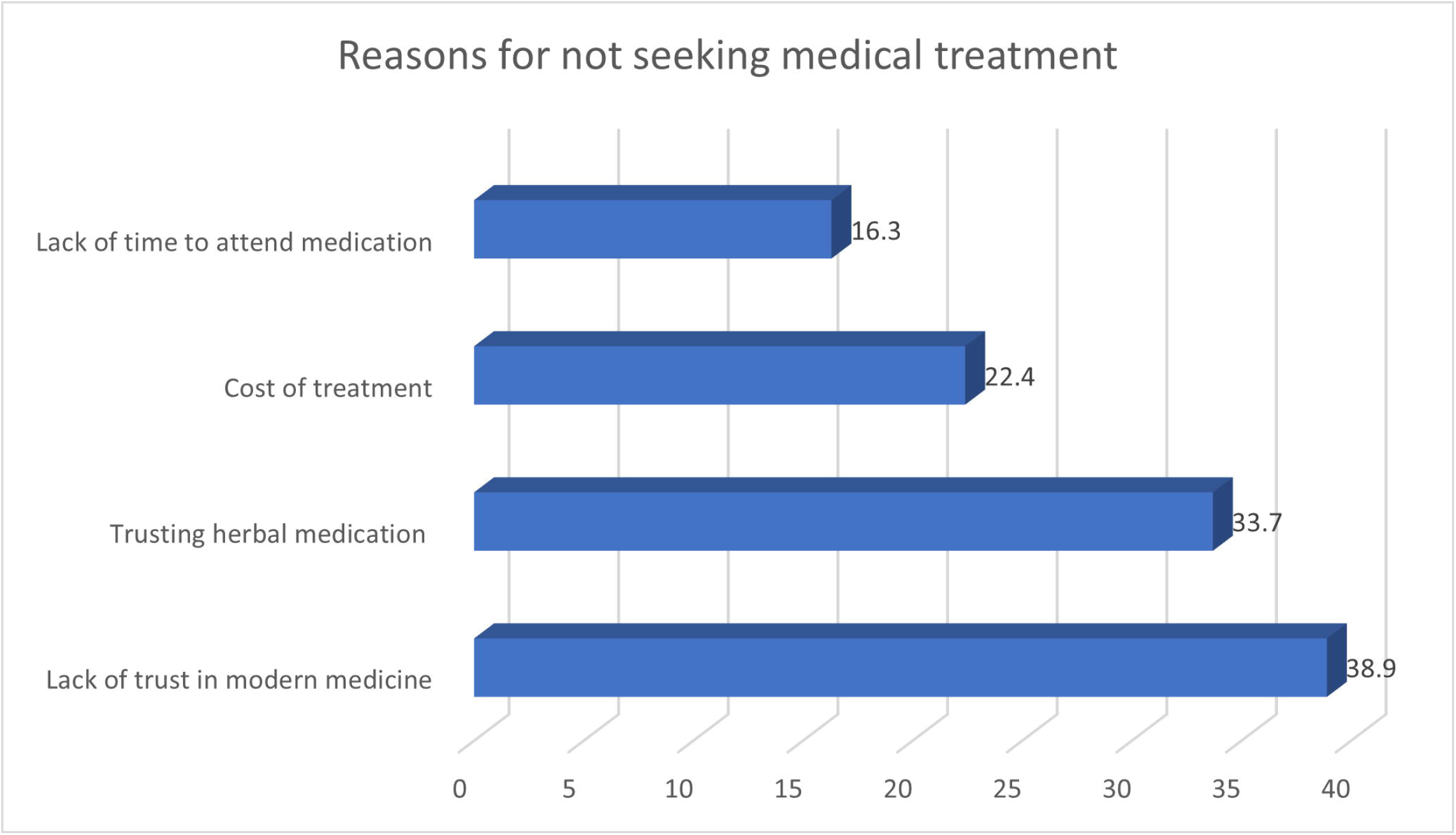
Reasons for not seeking medical treatment. In this study, lack of awareness 171 (40.3%), insufficient budget 114 (26.9%), and dependency on herbal medicine 99 (23.3%) were major constraints mentioned by the informants for not being able to control the disease within the community. Lack of such awareness creation programmes can also be explained by the fact 248 (58.6%) of the respondent confess that they do not think they are well informed about CL. Never having a chance to learn about the disease and inadequate source of information in the locality were mentioned by 203 (47.9%) of the respondents as reasons for not being well informed about the disease.

In this study, majority (64.6%, n=274) see CL as a very serious problem in their locality. Generally, 226 (53.2%) of the participants have a favourable attitude while 197 (46.4%) have unfavourable attitudes.

### Factors affecting the attitude of the community towards cutaneous leishmaniasis

In bivariate analysis, age (p< 0.048 and P< 0.018), occupation (p<0.028), place of birth (p< 0.017), and knowing CL infected person (p< 0.000) were seen significantly associated with attitude (Table 6). Therefore, those with age group of 24.5-34.5[COR=2.553; 95% CI, 1.007-6.475] and 34.5-44.5 [COR= 3.000; 95% CI, 1.208-7.448] have a favourable attitude about CL compared to those in the 18.0-24.5 age group.

**Table 6:**
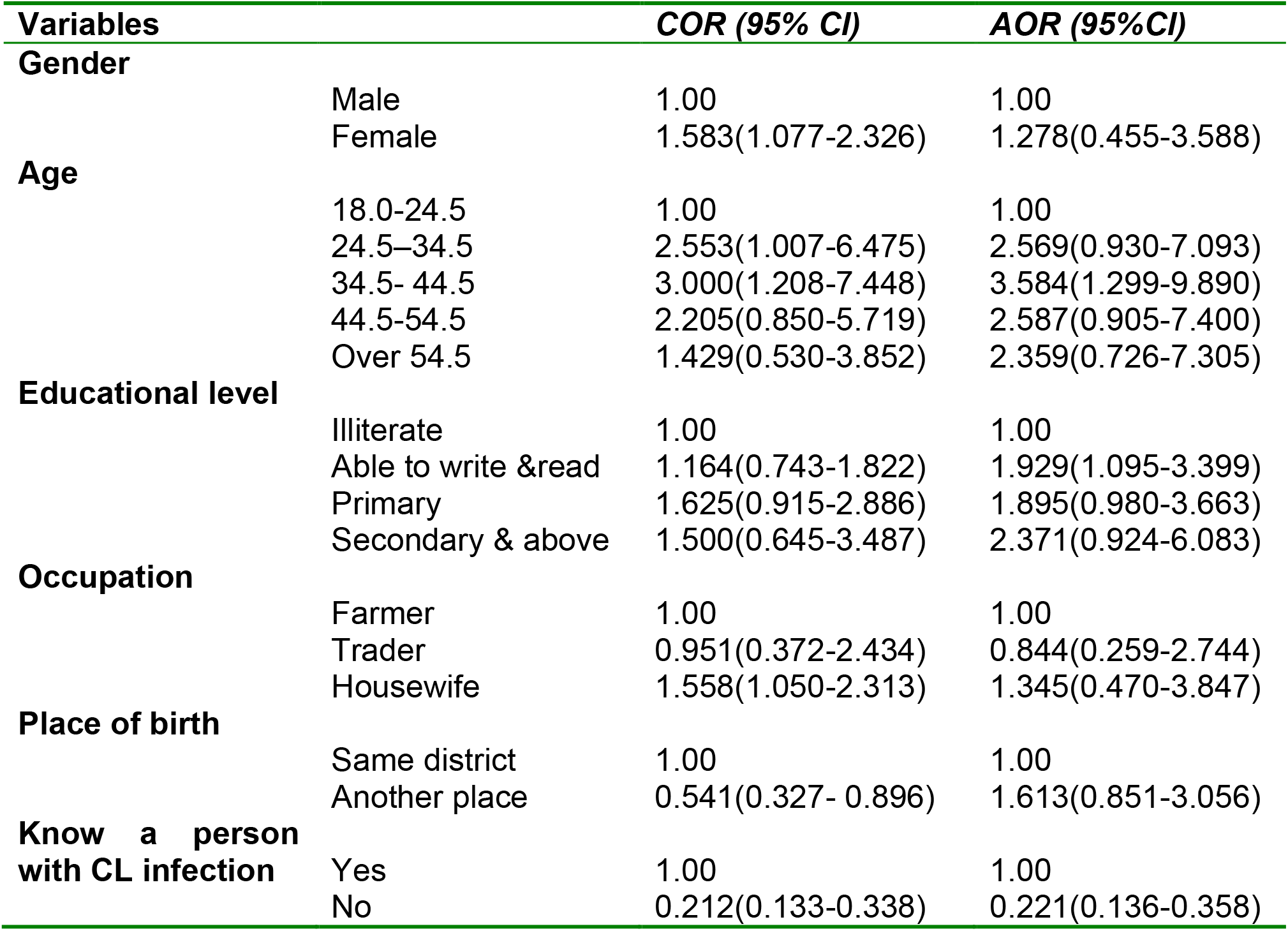
Comparison of socio-demographic variables with the attitude to CL (N=423).

| **Variables** |  | **COR (95% CI)** | **AOR (95%CI)** |
| --- | --- | --- | --- |
| **Gender** |  |  |  |
|  | Male | 1.00 | 1.00 |
|  | Female | 1.583(1.077-2.326) | 1.278(0.455-3.588) |
| **Age** |  |  |  |
|  | 18.0-24.5 | 1.00 | 1.00 |
|  | 24.5–34.5 | 2.553(1.007-6.475) | 2.569(0.930-7.093) |
|  | 34.5- 44.5 | 3.000(1.208-7.448) | 3.584(1.299-9.890) |
|  | 44.5-54.5 | 2.205(0.850-5.719) | 2.587(0.905-7.400) |
|  | Over 54.5 | 1.429(0.530-3.852) | 2.359(0.726-7.305) |
| **Educational level** |  |  |  |
|  | Illiterate | 1.00 | 1.00 |
|  | Able to write &read | 1.164(0.743-1.822) | 1.929(1.095-3.399) |
|  | Primary | 1.625(0.915-2.886) | 1.895(0.980-3.663) |
|  | Secondary & above | 1.500(0.645-3.487) | 2.371(0.924-6.083) |
| **Occupation** |  |  |  |
|  | Farmer | 1.00 | 1.00 |
|  | Trader | 0.951(0.372-2.434) | 0.844(0.259-2.744) |
|  | Housewife | 1.558(1.050-2.313) | 1.345(0.470-3.847) |
| **Place of birth** |  |  |  |
|  | Same district | 1.00 | 1.00 |
|  | Another place | 0.541(0.327- 0.896) | 1.613(0.851-3.056) |
| **Know a person with CL infection** |  |  |  |
|  | Yes | 1.00 | 1.00 |
|  | No | 0.212(0.133-0.338) | 0.221(0.136-0.358) |

The multiple logistic regression analysis shows that respondents who are in the 34.5 to

44.5 age group [AOR= 3.584; 95% CI, 1.299-9.890] and who can read and write [AOR= 1.929; 95% CI, 1.095-3.399] have favourable attitude than those who are in 18.0-24.5 age group and who are illiterate.

### Prevention practice of Sodo community regarding cutaneous leishmaniasis

When asked how CL was treated, the application of herbal remedies 222 (73.8%) and the use of specific drugs at health centre 133 (44.2%) were mentioned more frequently (Table 7).

**Table 7:** Practice of the Sodo community against CL (N=423).

| Variables<br>(multiple responses) |  | Frequency | Percentage |
| --- | --- | --- | --- |
| How CL was treated* | Herbal | 222 | 73.8 |
|  | Praying | 23 | 7.6 |
|  | Drugs given at health center | 133 | 44.2 |
|  | Not treated | 9 | 3.0 |
|  | Holy water | 19 | 6.3 |
|  | I do not know | 38 | 12.6 |
| Use preventable measure | Yes | 235 | 55.6 |
|  | No | 130 | 30.7 |
|  | I do not know | 44 | 10.4 |
| Having bed net in the house | Yes | 288 | 68.1 |
|  | No | 124 | 29.3 |
|  | Do not know | 11 | 2.6 |
| Sleeping outside or spending time outside | Yes | 19 | 4.5 |
|  | No | 392 | 92.7 |
| Using bed net when sleeping | Yes | 251 | 59.3 |
|  | No | 135 | 31.9 |
|  | Do not know | 36 | 8.5 |
| Using repellents | Yes | 128 | 30.3 |
|  | No | 244 | 57.7 |
|  | Do not know | 51 | 12.0 |
| Work time preference at high T° | Day time | 148 | 35.0 |
|  | Night | 37 | 8.7 |
|  | Both | 165 | 39.0 |
|  | Very early morning | 44 | 10.4 |
| Practice (overall) | Good | 213 | 50.4 |
|  | Poor | 201 | 46.9 |

\* N=301 number of individual who know someone infected with CL

About half 235 (55.6%) of participants in this study use preventive measures. The preventive measures used by participants in this study are bed nets 178 (42.1%), cleanliness 118 (27.9%), early detection and treatment 114 (27.0%), and insecticide 94 (22.2%) for prevention of the disease (Fig 4). Poor prevention practice was observed on personal protective measures (wearing covering clothes 5 (1.2%), use of insect repellents 28 (6.6%)) which are important as a first-line defence mechanism against sandfly bite.

**Fig 4:**
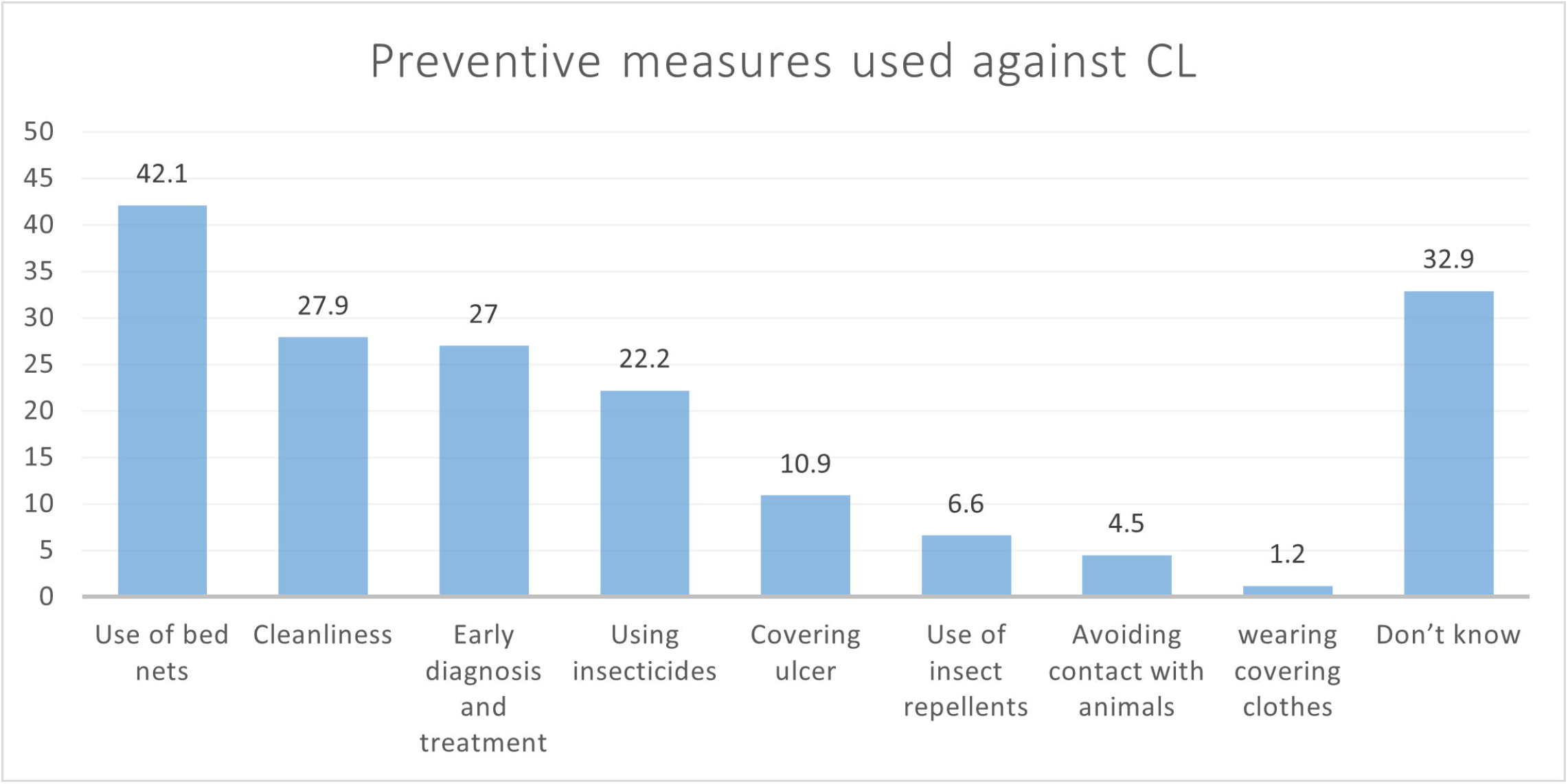
Type of major CL preventive measures used in Sodo District.

In this study, it was found that 251 (59.3%) use bed nets when they are sleeping. Looking into the number of bed nets in the current study, of those who use bed nets 105 (24.8%) and 69 (16.3%) have a maximum of two or three bed nets in the house, respectively. In this study, only 19 (4.5%) have a custom of spending time outside or sleeping outdoors.

More than half (57.7%) of the respondents do not use a repellent. Any type of insecticide has never been sprayed in 210 (49.5%) of the participants’ houses. Of the 170 (40.0%) insecticide sprayed houses, spraying was for protection against malaria. When asked about their preferred time of work at peak temperature, the respondents prefer to work at daytime 148 (35.0%) and both at daytime and night (39.0%). Generally, 210 (49.4%) of the respondent have good prevention practice while 214 (50.4%) of the respondents have poor practice.

### Factors affecting the practice of cutaneous leishmaniasis

Table 8 presents the result of logistic regression of demographic variables and overall practice score. Accordingly, in bivariate logistic regression female sex (p< 0.000), over 54 age group (p<0.024), being able to read and write (p< 0.030), housewives (p< 0.000), coming from other districts (p<0.003) and knowing someone with the diseases (p<0.000) are significantly associated with good practice.

**Table 8:** Socio-demographic characters and practice of participants (N=423).

| Variables |  | COR (95% CI) | AOR (95%CI) |
| --- | --- | --- | --- |
| **Gender** |  |  |  |
|  | Male | 1.00 | 1.00 |
|  | Female | 2.355(1.594- 3.480) | 0.841(0.309-2.292) |
| **Age** |  |  |  |
|  | 18.0-24.5 | 1.00 | 1.00 |
|  | 24.5–34.5 | 0.929(0.373-2.314) | 0.768(0.293- 2.019) |
|  | 34.5- 44.5 | 0.498(0.205- 1.208) | 0.453(0.174- 1.181) |
|  | 44.5-54.5 | 0.695(0.273- 1.767) | 0.703(0.258- 1.914) |
|  | Over 54.5 | 0.323(0.121- 0.863) | 0.359(0.123- 1.048) |
| **Educational level** |  |  |  |
|  | Illiterate | 1.00 | 1.00 |
|  | Able to write & read | 0.605(0.384-0.952) | 0.852(0.493- 1.475) |
|  | Primary | 1.556(0.873- 2.774) | 1.797(0.938- 3.444) |
|  | Secondary & above | 0.703(0.305- 1.628) | 0.776(0.309-1.948) |
| **Occupation** |  |  |  |
|  | Farmer | 1.00 | 1.00 |
|  | Trader | 1.360(0.531- 3.486) | 1.345(0.423-4.277) |
|  | Housewife | 2.548(1.705- 3.809) | 2.142(0.768- 5.972) |
| **Place of birth** |  |  |  |
|  | Same district | 1.00 | 1.00 |
|  | Another place | 2.153(1.300- 3.567) | 1.335(0.718- 2.483) |
| **Knowing someone with CL** |  |  |  |
|  | Yes | 1.00 | 1.00 |
|  | No | 0.300(0.191- 0.472) | 0.315(0.195- 0.510) |

As a result, female [COR=1.5; 95% CI, 1.031-2.233], those who can read and write [COR=1.5; 95% CI, 1.031-2.233], housewives [COR=1.5; 95% CI, 1.031-2.233], those who originally come from other districts [COR=1.5; 95% CI, 1.031-2.233] and those who know someone infected with the disease have good practice than men. In contrast, illiterates, farmers, those who are born in Sodo and those who previously did not meet with a person infected with the disease.

In multiple logistic regression only knowing someone infected with the disease show statistical significance relation with good practice regarding CL [OR=0.315; 95% CI, 0.195-0.510)].

### Overview of overall knowledge attitude and practice

Regarding the overall knowledge of the Sodo community about CL, 263 (61.9%) have satisfactory knowledge of CL while 161 (38.1%) have unsatisfactory knowledge. The percentages of study participants showing favorable and unfavorable attitudes about CL are 53.4% (n=226) and 46.6% (n=197) respectively. About half 213 (50.4%) of the participants exhibit good preventive practice against the disease while the rest 210 (49.6%) have a poor practice of CL (Fig 5).

**Fig 5:**
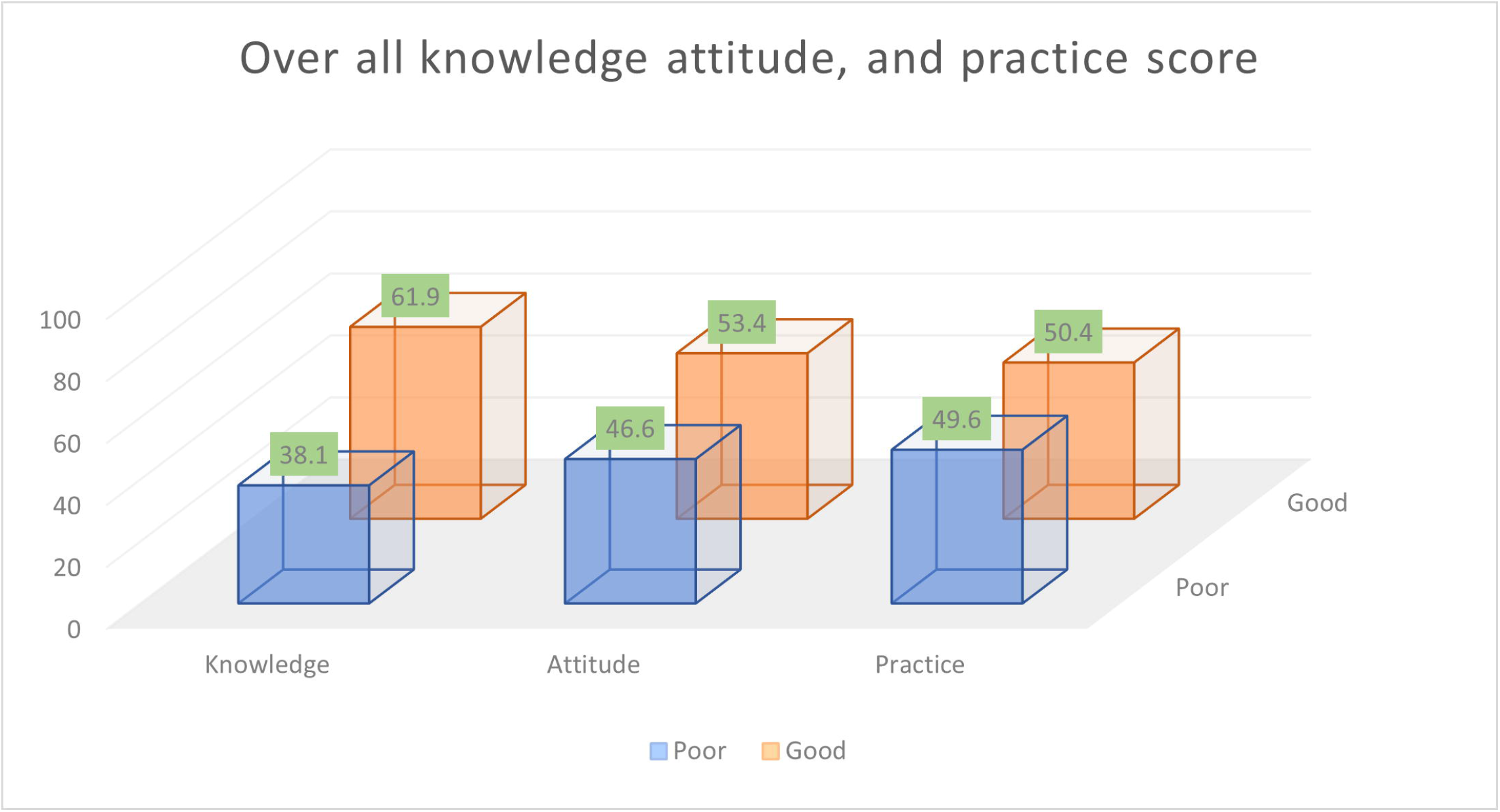
Overall KAP score of the community.

Comparing overall practice with knowledge and attitude in the current study (Table 4.11), the findings indicate that 262 (61.9%) respondents with satisfactory knowledge 173 (40.9%) have a good practice. The association between knowledge and practice was statistically significant at p<0.000. Accordingly, the odds of having a good practice are nearly five times higher for those who have satisfactory knowledge than those who have unsatisfactory knowledge.

**Table 4.11:** Comparison of overall knowledge, attitude, and practice (N=423).

|  | PRACTICE |  | Total No (%) | p- value | COR (95% CI) |
| --- | --- | --- | --- | --- | --- |
|  | Poor<br>No (%) | Good<br>No (%) |  |  |  |
| **Knowledge** |  |  |  |  |  |
| Unsatisfactory | 121(28.6) | 40(9.5) | 161(38.1) | 1 |  |
| Satisfactory | 89(21.0) | 173(40.9) | 262(61.9) | 0.000 | 4.722(2.992 - 7.454) |
| **Total** | 210(49.6) | 213(50.4) | 423(100) |  |  |
| **Attitude** |  |  |  |  |  |
| Unfavourable | 134(31.7) | 63(14.9) | 197(46.6) | 1 |  |
| Favourable | 76(18.0) | 150(35.5) | 226(53.4) | 0.000 | 3.213(2.081 - 4.960) |
| **Total** | 210(49.6) | 213(50.4) | 423(100) |  |  |

A similar relationship was seen between attitude and practice as of the 226 (53.4%) participants who have favorable attitude, 150 (35.5%) also have a good practice. This was also statistically significant at p<0.000 with individuals with favourable attitude having three times more likely to have a good practice.

## DISCUSSION

In the current study, 216 (51.1%) of the participants identified case of CL after shown a picture illustrating typical lesion of CL reported from Ethiopia and 350 (82.7%) of the participants have heard about CL. This was higher compared to reports from India (39.8%) (16), West Alexandri (10%) (17), Paraguay (29.0%) (18) and Southwestern Iran (47.9%) (19). Higher percentage reports of CL lesions awareness such as 75.0%, 82.0%, and 85.9% were seen from Nigeria among physicians in health care facilities (20), from communities in Ghana (21) and communities in Colombia respectively (22). Even majority (91.7%) of the indigenous community in French Guiana and Brazil were able to recognise a photograph with classic case of CL from French Guiana (23). The difference between these records and the current study may be because these studies were conducted among individuals who are health professionals (Nigeria) and among people during CL occurrence were highest (5) in the country (Colombia).

In the present study, majority of study population 350 (82.7%) have heard about CL. This was in line with a study from Southern Iran (24). The finding in the current study was greater than the findings from Ethiopia (67.6%) (25), Paraguayan (71.0%) (26). However, it is lower than those from other research, which show that 97.3% and 92.2% of participants in Sri Lanka and Iran, respectively, have heard about CL before (27). This disparity could be because the other studies were conducted among health professionals (in Sri Lanka) and the difference in educational status of mothers of CL-affected children in Iran (where only 6.2% are illiterate) and the current study (where only 6.2% are illiterate) (where 51.1% are illiterate).

In the current study, 206 (53.4%) of the respondents do not know the cause of CL. Another 252 (59.4%) do not know ways of disease transmission. Contrary to this, higher proportion of participants (80.0%) in Ecuador knew how CL is transmitted (28). Similarly, in Southern Iran, more than half of the respondents informed that microbe cause CL, and even a few 1.2% name Leishmania parasite as the causal agent of CL (24). In Sri Lanka 97.75% of the study population have good understanding of CL transmission (27).

The current study revealed that plaque and pus-containing wounds were mentioned as the main clinical symptom of CL by 268 (63.4%) and 206 (48.6%) of the respondents, respectively. Moreover, 92 (21.7 %) confess that they are not aware about any symptoms of CL. Knowledge of the symptoms and indicators of CL can aid individuals and their families in avoiding the disease (29). Skin infection, fever, liver and spleen enlargement, and anaemia are the most common leishmaniasis symptoms in Pakistan, according to 33.6%, but the majority (56.0%) are uninformed (3). According to a study conducted in Ethiopia, 59.6% of participants recognised swollen legs as a prominent indication, whereas 11.0% were unable to name any sign (25). The majority (95.8%) of students at Airbase Isfahan knowledgeable about signs and symptoms of CL (30). In Guatemala, 96.7% of the 425 heads of families asked could appropriately describe a typical CL lesion (31).

In this study concerning knowledge of the participants about the sandfly vector, nearly half 210 (49.6%) reported to know the biting and blood-sucking behaviour after being shown a picture of phlebotomine sandfly collected from nearby districts. These indicate a low level of awareness regarding the sandfly vector. The finding in the current study is in line with report from Tigray region of Ethiopia (32). No knowledge of the Leishmania vector was reported from Southwestern Iran (19), Pakistan (3). Preventive methods may fail due to such kind of knowledge gap, making education about this vector essential (33). Lack of information about CL and its vectors is an issue for implementing prevention measures and seeking early treatment (3).

The vector’s preferred breeding area and biting time were unknown to 123 (29.0%) and 139 (32.8%) of the participants in the current study, respectively. Sandflies are most active throughout the evening, nocturnal, and twilight hours (from dusk to dawn), and are less active during the hottest hours of the day. If they are disturbed, though, they will bite at any time (34). The most commonly stated breeding areas of the vector include dirty places (218 (51.5%), water ponds (194 (45.9%), and waste collection sites (111 (26.2%). Cervices in the house (18.3%), thatched roof (17.4%), and cattle shades (20%) all received a little percentage of the vote (4.7%). However, sand flies’ immature stages are more concentrated in microhabitats with specific characteristics, such as the presence of organic materials, wetness, and low light levels (35). In the same Pakistani survey, 14.0% of respondents believed that sandflies bite at dusk and down, followed by at any time of day (13.2%), during the daytime (10.0%), and at midnight (8.0%), while the majority (54.8%) were uninformed (3). Due to a lack of knowledge regarding sandfly biting times in endemic areas, personal protection measures such as mosquito nets, insecticides, and mosquito repellent may be used less frequently during those times. A case control study in Nepal demonstrated the effectiveness of employing bed nets for prevention in VL (36).

Regarding factors affecting knowledge of the study participants, the bivariate logistic regression shows that gender, occupation, education, location and previous knowledge of infected person are associated with knowledge about CL. None of these factors have significant association in multivariate logistic regression.

The Sodo community have favorable attitude about CL treatment as 61.7% believed that CL can be treated. This is important for seeking treatment and reduce the disease prevalence. A larger proportion (70.4%) of respondents in Pakistan thought that the disease is curable (3). In contrast, in Ethiopia, most participants (54.8%) believe CL is not treated, and 34.9% do not know if CL is treatable (25). Similarly, only 48.0% of people in Southern Iran believe CL can be treated (24). If people in a community have a positive attitude toward CL treatment, they are more likely to visit a healthcare professional for regular check-ups.

In this study, more than half 264 (62.4%) of the study subjects think CL infection causes disfiguring and 67 (15.8%) consider CL infection to result in both disfiguring and death while 54 (12.8%) have no idea on the outcome of CL infection. More than half (57.4%) of participants included in a similar study conducted in other parts of Ethiopia had the attitude that CL had a disfiguring outcome (25). Similarly, in Syria, appearance of the lesion followed by permanent skin mark was mentioned by most of the respondents (37).

In this study, more than half of the respondents (59.3%) chose indigenous medicine based on the use of herbs by traditional healers to treat CL. In a comparable research of Ethiopians’ treatment preferences, the vast majority [235 (59.9%)] believed that therapy from traditional healers was successful (25). The use of traditional home treatments was more frequent in Colombia (38) and French Guiana (23). However, the use of traditional remedies is necessity rather than a choice since biomedical treatments for leishmaniasis is not available (23). In Ethiopia, only very few health centres diagnose leishmaniasis therefore, most cases are handled traditionally using herbal remedies or applying heat with fire from charcoal (11).

Never having a chance to learn about the disease and inadequate source of information in the locality were mentioned by 203 (47.9%) of the respondents as reasons for not being well informed about the disease. A similar study indicates that as many as 97.3% of the participants had never had any formal education on leishmaniasis (21). In the current study, most of the study participants 329 (77.4%) are willing to take part if there would be any control and prevention programmes as 274 (64.6%) of the participants see CL as a very serious problem in their locality. Even if awareness creation and participation of communities in control interventions are vital, Ethiopia does not have a control program against CL though there is a guideline for the diagnosis, treatment, and prevention of CL since 2013 (11). Awareness creation campaigns are also rarely available to few (1.7%) local inhabitants in West Alexandria (17). Almost all, 375 (95.7%) of the study participants in Ethiopia, did not participate in the CL control activities (25). However, the availability of such control activities in the area was not mentioned in this study. So, this report may reflect the unavailability of designed control programmes rather than the low engagement of the community.

In this study, majority (64.6%, n=274) see CL as a very serious problem in their locality. Similarly, in Nigeria (20), and Paraguay (18), CL was regarded a serious public health problem in 87.0%, 53.0%, and 10.0% of communities, respectively (18). On the other hand, CL was not regarded as a serious disease in India by any of the respondents (16).

In the bivariate analysis, age (p< 0.048 and p< 0.018), occupation (p< 0.028), place of birth (p< 0.017), and knowledge of someone infected with CL (p< 0.000) show statistical significance value for a favourable attitude of CL.

The multiple logistic regression analysis shows that respondents who are in the 34.5-44.5 age group [AOR= 3.584; 95% CI, 1.299-9.890] and who can read and write [AOR= 1.929; 95% CI, 1.095-3.399] have a favourable attitude than those who are in 18.0-24.5 age group and who are illiterate. Similar reports were found in a study conducted in Iran with education being a stronger predictor for the attitude (39). Respondents with no history of knowing a person infected with the disease [AOR= 0.221; 95% CI, 0.136-0.358] are less likely to have a favourable attitude towards CL than those who previously know a person infected with CL.

Prevention practice of Sodo community regarding cutaneous leishmaniasis was assessed by eight questions. When asked how CL was treated for those who were exposed or know someone infected with CL, the application of herbal remedies (73.8%) and the use of specific drugs at health centre (44.2%) were mentioned more often. Half of the communities in Paraguay (57.14%) sought care from the hospital for CL/MCL (18). The use of herbal medication causes underreporting of the disease making the understanding of the true burden of the disease difficult. About half 235 (55.6%) of participants in this study use preventive measures. In Ecuador, many of the study participants (81.0%) had a better practice to CL. The difference in preventive practice between participants these studies may be the presence of vector control program in Ecuador. These will enhance awareness of the community hence leading to better preventive practices. In addition, notification of leishmaniasis is mandatory in Ecuador with passive and active detection of cases is in place (40). In contrast, there is no national control programme for leishmaniasis and sandfly vector in Ethiopia.

Participants in this study used bed nets 178 (42.1%), cleanliness 118 (27.9%), early detection and treatment 114 (27.0%), and insecticide 94 (22.2%) for prevention of the disease. Another study conducted in Ethiopia found that participants utilised bed nets (21.7%) and DDT (19.4%) as preventative strategies (25). In Iran, 16.1%, 14.8%, 3.7%, and 12.3% of the population had utilised medications, pesticide sprays, repellents, and bed nets to prevent the sickness, respectively (19). Admission to a hospital (31.2%), cleanliness (27.2%), usage of bed nets (26.0%), isolation of patients (9.2%), and dietary precaution (6.4%) were all cited as approaches for patient care with leishmaniasis in Pakistan (3). The use of a bed net (37.0%), insecticide (41.0%), hygiene (33.0%), adequate face and hand washing (19.0%), and bathing (16.5%) were all mentioned by participants in Southern Iran (24).

In the current study, poor prevention practice was observed on personal protective measures (wearing covering clothes 5 (1.2%), use of insect repellents 28 (6.6%) which are important as a first line defense mechanism against sandfly bite. The best way to prevent infection is to protect from sandfly bites by following these preventive measures to decrease the risk of being bitten. The use of personal protection is still considered the first line of prevention for leishmaniasis in endemic areas because sandfly eradication is difficult, chemoprophylaxis drugs are not effective and vaccines are still under development (26). Studies report that most of the participants did not use protective methods even though they know the importance of vector control as a key to prevent vector-borne diseases (41). The proportion of participants wearing protective clothing and using repellent (18.0%) as their ways of prevention was low in Ghana (21). In French Guiana, methods of leishmaniasis prevention for 36.4% of the population was mainly based on the topical application of carapa seed oil mixed with annatto seed paste as repellents (23).

In this study, it was found that 251 (59.3%) use bed net when they are sleeping. With regards to the prevention of CL in the endemic communities, 39.6% of the participants reported that they always use bed net (Doe et al. 2019). The majority (85.0%) of patients with post kalaazar dermal leishmaniasis in India use bed net while sleeping (42). On the contrary, bed net use was practiced by small proportion of the community in West Alexandria (17) and Pakistan (3). In Paraguay, most of the population 86.0% (397/463) never used a bed net (18). Bed nets provide a safe haven for those who sleep beneath them. Bed nets treated with pesticides, on the other hand, provide superior protection than untreated nets since the insecticides used to treat bed nets kill insects. The pesticides also repel vectors, which reduces the number of insects that enter the residence and try to feed on the people’ blood (43). The malaria prevention and control programme in Ethiopia targets 100.0% household coverage with two ITNs per household in all malaria endemic areas (44). However, there is a geographical difference in the distribution of malaria and leishmaniasis in the country. Therefore, though efforts made to prevent malaria also reduce the burden of leishmaniasis in some areas, a specifically designed national leishmaniasis control programme is still in need.

Only 19 (4.5%) of those polled have a habit of spending time or sleeping outside. Another survey conducted in southern Ethiopia found that most respondents (59.2%) did not sleep outside (25). Different sleeping habits were seen in the northern region of the country, where more than half of the population, 166 (62.88%), had experienced outdoor sleeping, particularly during hot weather (32). The disparity is due to the temperature and humidity differences between the southern and northern parts of the country.

In this study, more than half of the respondents (244 (57.7%) do not use a repellant. Among the Visceral Leishmaniasis prevention methods used in Western Tigray, Ethiopia, 88 (33.33%) and 163 (61.74%) respondents employ repellents and chopped plant parts, respectively, to protect oneself from biting flies (32). In order to avoid insect-borne infections, the Centers for Disease Control and Prevention (CDC) suggests using insect repellents. Human breath (carbon dioxide) and skin smells attract mosquitoes and other blood-feeding insects. Repellents are one of the personal protection measures that contain a chemical that makes the person undesirable to the vector, preventing biting (45).

When asked about their preferred time of work at peak temperature, respondents prefer to work at daytime 148 (35.0%) and both at daytime and night (39.0%). Another study conducted in Ethiopia found that the majority of the study participants worked on their farms during the day [375 (95.7%)] (25). Since a sandfly bite at night, working in these times pose exposure to the bite and disease transmission.

In multiple logistic regression, only knowing someone infected with the disease shows statistical significance relation with good practice regarding CL [OR=0.315; 95% CI, 0.195-0.510]. Similarly, a family history of CL was a stronger predictor for the practice in Iran(39).

Regarding the overall Knowledge of the Sodo community regarding CL, 263 (61.9%) have satisfactory knowledge of CL. Similar reports were seen from the southern part of Ethiopia where (67.6%, n=265) study participants were knowledgeable (25). In west Alexandria, Egypt, among the 20 primary health care physicians, 55% of the physicians had satisfactory knowledge (17). In Colombia, only 15% of homeowners had a “very excellent” understanding of CL, compared to 56.3% who had a “bad” or “poor” understanding of CL, according to these research (22). A survey conducted in Iran also shown that people in Iran had a poor understanding of CL (39). According to another study conducted in Iran, 211 people (52.1%) were unaware about CL(19). The discrepancy in the community awareness report in the aforementioned research can be explained by the study population and study period differences.

In the current study, the percentages of study participants showing favourable and unfavourable attitudes about CL are 53.4% and 46.6 % respectively. A similar study in Ethiopia revealed a lower percentage of positive attitude among 162 (41.3%) (25). A higher percentage (85%) of a good attitude score was seen among Egypt health care physicians showed a good attitude about leishmaniasis (17). Health care physicians would probably have better disease awareness owing to educational preparation and clinical experience. In Iran, average scores of attitudes were reported among households (39) and mothers of children with CL (33).

About half 213 (50.4%) of the participants in this study show good preventive practice against the disease while the rest 210 (49.6%) have a poor practice of CL. Mothers of children with CL have better practice compared to participants in this study as only 32.5% of the mothers had weak performance (33). This may be because females have a better attitude than males as shown previously in this study. However, the practice score of participants in this study is higher than reports from southern Ethiopia by Kebede et al.(25) where 62.5% of the population has poor practice. This may be because of differences in the study period. With the current study conducted recently, communities may have better access to knowledge. Students attending Isfahan’s Shahid Babaie Airbase’s middle and high schools were likewise found to have a lack of CL practice (30). This may be owing to the age factor as students in school are younger than populations in randomly selected communities.

Comparing overall practice with knowledge and attitude in the current study, the result indicates of 262 (61.9%) respondents with satisfactory knowledge 173 (40.9%) have a good practice. The association between knowledge and practice was statistically significant at p < 0.000 accordingly the odds of having a good practice are 5 times higher for those who have satisfactory knowledge than those who have unsatisfactory knowledge. A similar relationship was seen between attitude and practice as of 226 (53.4%) participants who have favourable attitude, 150 (35.5%) also have a good practice. Individuals with a positive attitude were three times more likely to have a good practice, which was statistically significant at P < 0.000. Both knowledge and attitude had a strong connection with practice (p-value 0.001) in a KAP study of Iranian homes (39). This indicates the effectiveness of educational activities in endemic areas for disease prevention. Practice efforts to seek and get health-related knowledge are linked to more health-oriented thinking and, in general, healthier behaviors (46).

## Conclusion

A satisfactory level of general knowledge was seen among 262 (61.9%) of the respondents. This result may not be considered enough for the prevention of the disease because one-third of the community has unsatisfactory knowledge. Considering the endemicity of the disease in the area, this level of knowledge is deficient especially when there is a knowledge gap on the cause and the vector of the disease which could be the most important knowledge to prevent the disease. Even though the community can recognise CL, transmission methods are poorly understood. This knowledge gap may affect the disease incidence and pose a risk for the endemicity.

The percentage of study participants showing favourable and unfavourable attitudes about CL is 53.4% (n=226) and 46.6 % (n=197) respectively. This compared to other findings is relatively higher. However, there is a concern on attitude about treatment choice of the community as the preferred choice of medication was the application of traditionally known herbs for 251 (59.3%) of the population.

About half 213 (50.4%) of the participants exhibit good preventive practice against the disease while the rest 210 (49.6%) have a poor practice of CL. There is also a bridge in the preventive practice of the community because personal protective measures, use of repellents and application of insecticides are low in addition to the predominant use of herbal remedies for treating the disease.

From the KAP survey results, evidence of discrepancies between knowledge attitude and practice is observed. About half (51.1%) of them were able to correctly name the disease when shown a picture with a typical case. However, more than half do not know the cause (53.4%) of the disease, 59.4% do not know how the disease is transmitted and 50.4% have no idea about the importance of the vector in disease transmission. Even though 82.7% heard about the disease, more than half (58.6%) think that they are not well informed about CL. Concerning the attitude of respondents about the disease, majority (77.5%) are willing to participate in CL prevention/control programmes and 64.5% think that the disease is very serious in their locality. Nonetheless, only 55.6% have a practice of using preventive measures against the disease.

Recommendations derived from the study findings are: Awareness of the community needs to be improved especially on cause and transmission means of the disease. Comprehensive effort must be exerted to increase community awareness on the importance of sandfly vector including breeding places, resting sites, and preferred biting time. The perception of the community also needs to be transformed because they have a low habit of utilisation of modern medication. This has to be coupled with provision of readily available CL treatment to the community. The community needs to be sensitised about the use of preventive measures especially personal protective measures that could create a barrier to sandfly bite, use of repellents and insecticide application. Community outreach on CL will have a huge advantage as it has been shown from the findings of these research those who know someone infected with CL proven to have better knowledge, attitude, and practice. The above-recommended awareness creation programmes should specifically target individuals of different ages and levels of education.

## Supporting information

Supplemental questioner

Supplemental picture

## Data Availability

Given our ethical clearance protocols, we cannot make the data available to other researchers as the consent forms explain to study participants that information we collect through this study endeavor private and the information will not be accessible to anyone else.

## Supporting information

S1Text. Knowledge Attitude and Practice survey questioner.

S2 Text. Picture shown to participants illustrating cases of CL reported from different parts of Ethiopia.

S3 Text. Picture of the vector that is believed to exist in the southern part of Ethiopia which was shown to the respondents.

## Acknowledgments

We would like to express our heartfelt gratitude to Sebseba Kebede for assisting with field operations, as well as the data collectors who actively participated in the study, shared their experiences, and contributed to the study’s success. We would also like to thank the study members who take part in the study.

