## Supplemental questioner for "Knowledge Attitude and Practice towards Cutaneous Leishmaniasis in Sodo District Southern Ethiopia"

**Supplementary information (S1)**

**KAP survey questioner (English version)**

**KAP SURVEY QUESTIONNAIRE**

**Survey objective:** To explore CL- related knowledge, attitudes and practice of persons bitten by suspected rabid animals

*Check one:* Date: ___ / ___ / ___

Baseline data collection; or	Sub city: _______ site: __________

Follow-up data collection	code: ________

*Information to read to respondent:*

We'd like to learn more about your CL knowledge, attitudes, and practices. We intend to identify your requirements and the most effective manner to deliver information to you, as well as any barriers to getting medical care. CL control will be improved with the information you supply. Your responses will be kept private and will not be shared with anybody. Your name will not appear on the questionnaire or be recorded in any other way. Your participation in the interview is entirely voluntary, and you may terminate it at any time.

Thank you for your assistance.

**Interviewer:** Place an X in the box of the selected answer(s).

Do not read responses unless the directions indicate.

A. **DEMOGRAPHIC INFORMATION**

**1.** How old are you? _____________ Years**.**

1. □ 15-24	3. □35- 44
2. □ 25–34	4. □ 45-54	□Over 55

**2.** What is your gender? (Record by observing)

1. □Male	2. □Female

3. What is the highest level of education you have completed?

1. □ No formal school	4. □College

2. □ Elementary or junior5. □Higher education (professional or post-graduate)

3. □High school (9-12grade)	6. □Religious schooling only

4. Duration of stay at residence ________________________________

5. Household number ___________________________________

6. Place of birth __________________________________

7. If migrant; Place of origin _______________ year of arrival ____________

8. Occupation ________________________________

1. □ Agricultural labor	2. □ Services

3. □ Skilled laborer	4. □Business

5. □government employee	7. □ Other (specify): _____________

8.1. If your income source is farming, the location of the farmland is

1= near home

2= near the river/________ (in Butajirra)

3 = both

9. Religion ___________________

1. □ Orthodox	5. □ Pagan

2. □ Muslim	6. □ Other (specify): _____________

3. □Protestant

4. □catholic

**B.** **GENERAL KNOWLEDGE OF CL**

10. Ask the respondent if they could name the disease after showing a picture of CL manifestation

1. □ Able to identify as CL	2. □ Unable to identify

3. □ other: ______________________

11. Have you heard about Cutaneous Leishmaniasis (Chewi)?1. □yes	2. □No

12. If your answer is ‘yes” to the question above, how did you hear about CL? (Please choose the three most effective sources.)

1. □ Newspapers and magazines	2. □TV	3. □Radio

4. □Billboards	5. □Health workers	6. □ Religious leaders

7. □Brochures, posters and other printed materials	8. □Teachers

9. □Family, friends, neighbors and colleagues

10. □Other (please explain): ___________________________________

13. What is the cause of CL?

1.□spiritual or hereditary	2. □ contact with patient

3. □virus	4. □Leishmanaia

5.□Mosquito bite	6. □the bite of sandfly

7. □Germ	8. □microbe

9. □I do not know

10. □other: __________________________________________________

14. What are the signs of CL?

1. □ fever with chills	3. □Plaque

2. □Skin rash	4. □Papule

5. □ Ulcer	6. □I do not know

7. □other: _____________________________________________________

15. Where in the body are lesions/scars of CL Located?

1. □ Forehead	2. □ Face	3. □ Nostril

4. □ Arm	5. □ Leg	6. □ Ear

8. □ Mixed	7. □I do not know

9. □ other: ____________________________________

16. Is CL an infectious disease/ can be transmitted from one person to another?

1. □yes	2. □No	3. □I do not know

17. Is there Possibility of acquiring leishmaniasis in travelling to endemic areas?

1. □yes	2. □no	3. □I do not know

18. Is complete cure from CL possible?

1. □yes	2. □no	3. □I do not know

19. Ask the respondent if they know about the biting and blood sucking behavior after showing live sand flies collected from their environment:

1. □ yes, I know	2. □ No, I don’t know

20. Where do you think is the breeding place of the vector?

1. □ Dirty place	4. □Damp and dark places

2. □ Cervices in the house	5. □ water ponds

3. □ Thatched roof	6. □ Garbage collection sites

7. □Cattle sheds

8. □ do not know

9. □Others: ______________________

21. When is the preferred biting time of the vector?
1. □Dusk	3. □Midnight
2. □anytime whether day or night	4. □ I do not know

C**. THE ATTITUDES TOWARDS CL**

22. Do you think CL can be treated?

1. □ yes	2. □ no	3. □ I have no idea

24. Do you believe that the occurrence of CL in one member of the family affects the economy of the whole family?

1. □ yes	2. □ no	3. □ I have no idea

25. What do you think the outcome of CL if not treated?

1. □ death	2. □ disability	3. □ self cure

4. □ I have no idea	5. □Other: ________________

26. Do you believe that environmental health is important for prevention of transmission?

1. □ yes	2. □ no	3. □ I have no idea

27. What do you think are the major constraints to control CL?

1. □an insufficient budget	3. □ Religious taboo

2. □ Trained professionals	4. □ Lack of appropriate legislation

5. □Awareness	6. □Lack of proper coordination

7. □ I do not know

8. □Others: _________________________________________________

28. What is yours preferred drug of choice for treatment of CL?

1. □ Specific medicine	2. □Indigenous medicine

3. □ Do not know

4. □other: ________________________________________________

28. If you would not go to the health facility, what is the reason? (Please check all that apply.)

1. □ Not sure where to go	1. □Yes	0. □No

2. □Cost	1. □Yes	0. □No

3. □ Difficulties with transportation/distance to clinic	1. □Yes	0. □No

4. □Do not trust medical workers	1. □Yes	0. □No

5. □Do not like attitude of medical workers1. □Yes	0. □No

6. □Cannot leave work	1. □Yes	0. □No

7. □Don’t want to find out that something is wrong 1. □Yes	0. □No

8. □ preference to use herbal medication	1. □Yes	0. □No

9. □Other (please explain): _______________________________________

29. How expensive do you think CL diagnosis and treatment is in this country?

1. □ It is free of charge	2. □It is reasonably priced

3. □ It is somewhat/moderately expensive	4. □It is very expensive

5. □I have no idea

30. Do you feel well informed about CL?

1. □Yes	2. □ No

31. If your answer to the above question is “no” whose mistake is that?

1. □Your negligence

2. □ You never had the chance to learn about rabies

3. □there is inadequate source of information in your locality

4. □other: _____________________________________________

32. Have you ever participated in CL control activities?

1. □Yes	2. □No (reason: ___________)

33. How serious a problem do you think CL is in your locality? (Check one.)

1. □Very serious	2. □Ordinary	3. □Not very serious

4. □I don’t know

**D. PRACTICES REGARDING CL**

34. How can someone with CL be treated? (Check all that are mentioned.)

1. □Herbal remedies	1. □Yes	0. □No

2. □ Home rest without medicine	1. □Yes	0. □No

3. □Praying	1. □Yes	0. □No

4. □ Specific drugs given by health centre1. □Yes	0. □No

5. □ vaccination	1. □Yes	0. □No

6. □holy water	1. □Yes	0. □No

7. □Do not know

8. □Other: _______________________________________________

35. Do you think CL is preventable?

1. □ yes	2. □ no	3. □ I have no idea

36. What are the preventive/control measures against CL?

1. □ Using insecticides	1. □Yes	0. □No

2. □ Early diagnosis and treatments	1. □Yes	0. □ No

3. □ Using covered clothes	1. □Yes	0. □No

4. □ Use of bed nets	1. □Yes	0. □No

5. □ Use of insect repellents	1. □Yes	0. □No

6. □ Reduce outdoor activities	1. □Yes	0. □No

7. □ avoiding contact with animals	1. □Yes	0. □No

8. □ cleanliness	1. □Yes	0. □No

9. □covering of the ulcers

10. □Do not know

11. □Other: _______________________________________________

37. Do you have bed net in the house?

1. □yes (how many: ______________________)	2. □ no

38. Do you have custom of spending time outside in the night or sleeping outdoor?

1. □ yes	2. □ no

39. Do you use bed nets when sleeping? 1. □ yes	2. □ no

40. When is your Work time preference when the temperature is high?

1.□Day time	2. □Night	3. □Both

41. Do you use repellents?	1. □yes	2. □ no

42. Do you properly perform garbage disposal

1. □ yes (how often; ______________)	2. □ no

43. Has your house ever been sprayed?

1. □ yes (how often; _________________________________________)

2. □ no (reason: __________________________________________)

Thank you very much for participating in our survey.
