## Supplemental picture for "Knowledge Attitude and Practice towards Cutaneous Leishmaniasis in Sodo District Southern Ethiopia"

**Supplementary information (S3)**

Picture of the vector that is believed to exist in the southern part of Ethiopia which was shown to the respondents.

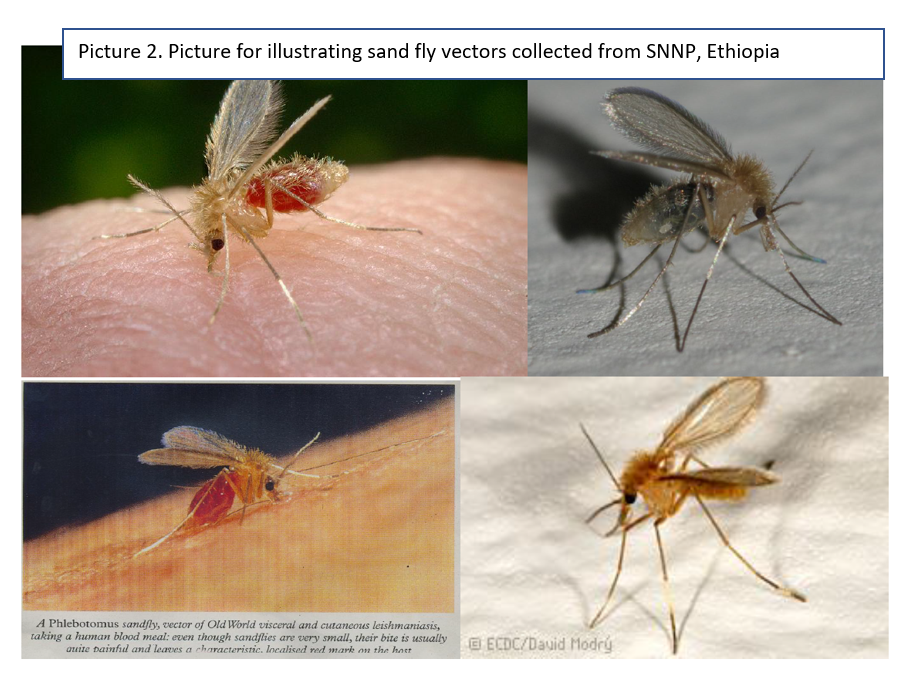
